# Effect of Menstrual Cycle and Menopause on Human Gastric Electrophysiology

**DOI:** 10.1101/2023.09.21.23295929

**Authors:** Alexandria H Lim, Chris Varghese, Gabrielle Sebaratnam, Gabriel Schamberg, Stefan Calder, Armen Gharibans, Christopher N Andrews, Daphne Foong, Vincent Ho, Shunichi Ishida, Yohsuke Imai, Michelle R Wise, Greg O’Grady

**Affiliations:** Department of Surgery, The University of Auckland, Auckland, New Zealand; Alimetry Ltd, New Zealand; Division of Gastroenterology, University of Calgary, Calgary, Alberta, Canada; Western Sydney University, Sydney, NSW, Australia; Graduate School of Engineering, Kobe University, Japan; Department of Obstetrics and Gynaecology, The University of Auckland, New Zealand

**Keywords:** electrophysiology, menstrual cycle, gastric physiology, body surface gastric mapping, motility

## Abstract

Chronic gastroduodenal symptoms disproportionately affect females of childbearing age; however, the effect of menstrual cycling on gastric electrophysiology is poorly defined. To establish the effect of the menstrual cycle on gastric electrophysiology, healthy subjects underwent non-invasive Body Surface Gastric Mapping (BSGM; 8×8 array), with validated symptom logging App (Gastric Alimetry^Ⓡ^, New Zealand). Participants were premenopausal females in follicular (n=26) and luteal phases (n=18). Postmenopausal females (n=30) and males (n=51) were controls. Principal gastric frequency (PGF), BMI-adjusted amplitude, Gastric Alimetry Rhythm Index (GA-RI), fasted-fed amplitude ratio (ff-AR), meal response curves, and symptom burden were analysed. Menstrual cycle-related electrophysiological changes were then transferred to an established anatomically-accurate computational gastric fluid dynamics model (meal viscosity 0.1 Pas), to predict the impact on gastric mixing and emptying. PGF was significantly higher in the luteal vs. follicular phase (mean 3.21 cpm, SD (0.17) vs. 2.94 cpm, SD (0.17), p<0.001) and vs. males (3.01 cpm, SD (0.2), p<0.001). In the computational model, this translated to 8.1% higher gastric mixing strength and 5.3% faster gastric emptying for luteal versus follicular phases. Postmenopausal females also exhibited higher PGF than females in the follicular phase (3.10 cpm, SD (0.24) vs. 2.94 cpm, SD (0.17), p=0.01), and higher BMI-adjusted amplitude (40.7 µV (33.02-52.58) vs. 29.6 µV (26.15-39.65), p<0.001), GA-RI (0.60 (0.48-0.73) vs. 0.43 (0.30-0.60), p=0.005), and ff-AR (2.51 (1.79-3.47) vs. 1.48 (1.21-2.17), p=0.001) than males. There were no differences in symptoms. These results define variations in gastric electrophysiology with regard to human menstrual cycling and menopause.

**New and Noteworthy:** This study evaluates gastric electrophysiology in relation to the menstrual cycle using a novel non-invasive high-resolution methodology, revealing substantial variations in gastric activity with menstrual cycling and menopause. Gastric slow wave frequency is significantly higher in the luteal versus follicular menstrual phase. Computational modelling predicts that this difference translates to higher rates of gastric mixing and emptying in the luteal phase, which is consistent with previous experimental data evaluating menstrual cycling effects on gastric emptying.

## Introduction

Chronic gastroduodenal symptoms are common, with an estimated global prevalence of 10.6% (1), but pathophysiology remains heterogeneous, contributing to difficulties in diagnosis and management (2). Young females are known to suffer from chronic nausea, vomiting, reflux, and epigastric pain at higher rates compared to the general population (1,3). As a result, they are more likely to experience reduced quality of life, higher mental health burdens, and accrue higher health economic costs (4,5).

Female sex hormones that fluctuate during the menstrual cycle are known to impact gastric emptying rates, and have been hypothesized to contribute to gastroduodenal symptoms (6–8). Elevated levels of progesterone may modulate gastric contractility, mediated through receptors found along the gastrointestinal (GI) tract (9–11) and may influence gastric dysrhythmia inducibility (9). However, changes in gastric electrophysiology occurring during the menstrual cycle remain unclear, due to a lack of detailed studies using accurate technologies (12).

Recent advances in BSGM (also termed high-resolution electrogastrography) now provide the ability to undertake reliable non-invasive assessments of gastric myoelectrical function in humans (13,14). This technique assesses both underlying gastric slow wave activity, generated by interstitial cells of Cajal, and associated gastric contractions (15–17). The aim of this study was therefore to evaluate the influence of the menstrual cycle on gastric function using BSGM, under the hypothesis that variation in gastric function would be observed with phases of the menstrual cycle and with menopause.

## Methods

This was a multi-centre, observational study of healthy participants conducted in Auckland (New Zealand), Western Sydney (Australia) and Calgary (Canada). Ethical approval was granted under the following references: AHREC 1330 (Auckland Health Research Ethical Committee), REB19-1925 (University of Calgary), H13541 (Human Research Ethics Committee, Western Sydney). All participants provided written informed consent.

### Study Population

Demographic data, including age, sex and BMI, were collected from all subjects. Healthy controls were eligible for inclusion if they were ≥ 18 years, not pregnant or breastfeeding, and did not experience any chronic gastroduodenal symptoms (nausea, vomiting, abdominal pain, early satiation) meeting Rome IV criteria for gastroduodenal disease (18). Male participants were included as a reference group. Subjects were excluded if they had an incomplete test record, poor test quality (artifact > 50% or poor impedance) or any of the following comorbidities: current gastrointestinal (GI) infection, history of inflammatory bowel disease (IBD), GI malignancy, previous GI surgery, regular cannabis use or metabolic or neurogenic disease known to affect gastric function. Specific exclusion criteria related to BSGM, per the Gastric Alimetry Instructions For Use, were BMI > 35 kg/m^2^, active abdominal wounds or abrasions, fragile skin, and adhesive allergies.

Female participants were excluded from menstrual cycle analysis if they did not experience periods, were perimenopausal (between the ages of 50 - 52) or had self-defined their menstrual cycles as ‘irregular’ at the time of BSGM testing. Participants on any form of local or systemic hormonal contraception were also excluded.

### Gastric Alimetry Body Surface Gastric Mapping

Participants were scheduled to wear the Gastric Alimetry device (a non-invasive gastric mapping device comprising a 64-electrode array and a wearable reader) for a total of 4.5 hours after an overnight fast. A standardised test protocol was performed (15), consisting of a 30-minute pre-prandial recording followed by a test meal (Ensure (230 kcal, 230 mL; Abbott Nutrition, Chicago, IL) and an oatmeal energy bar (Clif Bar Nutrition Bar Chocolate Chip; 250 kcal, 5 g fat, 45 g carbohydrate, 10 g protein, 7 g fibre; Clif Bar & Company, Emeryville, CA), or a similar dietary equivalent, consumed over ten minutes, followed by a 4-hour post-prandial recording. During the test, participants were asked to rate any upper gastrointestinal symptoms of upper gut pain, nausea, bloating, heartburn, stomach burn, and excessive fullness every 15 minutes on visual analogue scales from 0 to 10 (0 indicating no symptoms; 10 indicating the worst imaginable extent of symptoms) using the validated Gastric Alimetry App (19). These data were also used to calculate the validated ‘Total Symptom Burden Score’. Discrete events of vomiting, belching, and reflux were recorded (19). Further detail on the standard Gastric Alimetry BSGM protocol is described extensively in other publications (13,20).

### Body Surface Gastric Mapping Metrics

Four spectral metrics reported by the Gastric Alimetry system were employed (21): Principal Gastric Frequency (PGF; the sustained frequency associated with the most stable gastric slow wave oscillations; normative interval 2.65 - 3.35 cpm), BMI-adjusted amplitude (normative interval 22 - 70μV), the Gastric Alimetry Rhythm Index (GA-RI; quantifying the extent to which gastric electrical activity is concentrated within a narrow frequency band over time relative to the residual spectrum; normative interval ≥0.25), and Fed:Fasted Amplitude Ratio (ff-AR, normative interval ≥1.08). A detailed description and validation for each metric are detailed in the supplementary methods and elsewhere (21,22).

### Menstrual Cycle Phase Evaluation

Menstrual cycle data was captured in REDCap (Version 13.1.5, Vanderbilt University, Tennessee, USA) using a pre-specified questionnaire regarding contraceptive use, cycle regularity and date of last menstrual period.

The number of days between the start of the last menstrual period and the study date was calculated. An average 28-day cycle was assumed for all participants, and parameters for follicular and luteal phases were defined using an adaptation of the cycle-day-based model by Pitchers and Elliot-Sale (23). Participants with greater than 28 days since their last menstrual period were excluded. The onset of menses was considered Day 1, the follicular phase was defined as days 0-15, and the luteal phase was defined as days 16-28. Significant increases in progesterone levels have been reported only at 15 days after the start of the last menstrual period (24). The low-progesterone ovulatory phase described by Pitchers and Elliot-Sale (days 13-15) was amalgamated with the early and late follicular phases (days 1-5 and 6-12, respectively). Females over the age of 52 with no periods were considered post-menopausal. Perimenopausal females were excluded in accordance with the recommendations from Schmalenberger et al. (25).

### Statistical Analysis Methods

All analyses were performed using R software (Version 4.2.3, R Foundation for Statistical Computing, Vienna, Austria). BSGM metric and symptom comparisons were made using ANOVA tests in normally distributed data and Kruskall-Wallis and Wilcoxon signed-rank tests in non-normally distributed data for further pairwise testing. Analyses were considered statistically significant below the threshold of p < 0.05. Parametric data are presented as mean and standard deviation (SD). Non-parametric data are presented as the median and interquartile range (IQR).

### Computational Model

A previously established computational model of gastric fluid dynamics, mixing, and emptying was employed to estimate the physiological consequences of the defined electrophysiological changes across the menstrual cycle. A detailed description of the model setup is reported in prior publications (26–28). In brief, a three-dimensional anatomical model was established from a realistic gastric geometry, with peristaltic contractions modelled from corpus to terminal antrum based on MRI imaging studies (29). Wave frequency parameters were experimentally defined by the results in the present study, whereas wave velocity was varied according to defined relationships between frequency and velocity in the human stomach, with distinct values assigned to the corpus and proximal antrum versus distal antrum (28,30). Pyloric opening and closure was modelled according to established relationships with distal gastric contraction dynamics (26). Liquid food was employed with a viscosity of 0.1 Pas, modelled as an incompressible Newtonian fluid for simplicity. Free-surface flow modelling was used to incorporate the effects of gravity, and conservation laws of mass and momentum were solved numerically using the lattice Boltzmann method, under moving boundary conditions prescribed to the wall (refer to (27,28) for technical details). Separate gastric models were established for follicular and luteal menstrual phases.

## Results

Participants were recruited from New Zealand (96/121, 79%), Australia (12/121, 10%) and Canada (13/121, 11%). The majority of subjects were females (69/121, 58%). The median age was 40 years (range 18 - 78) for females and 32 years (range 19 - 75) for males. The mean body mass index (BMI) was 23.7 ± 3.3 kg/m^2^ (range: 16.6 to 33). Baseline demographic features were similar amongst participants in the follicular and luteal phases of their menstrual cycles, as detailed in **Table 1**. A diagram demonstrating study inclusion, exclusion, and resultant subgroups is provided in **Figure 1**.

**Figure 1:**
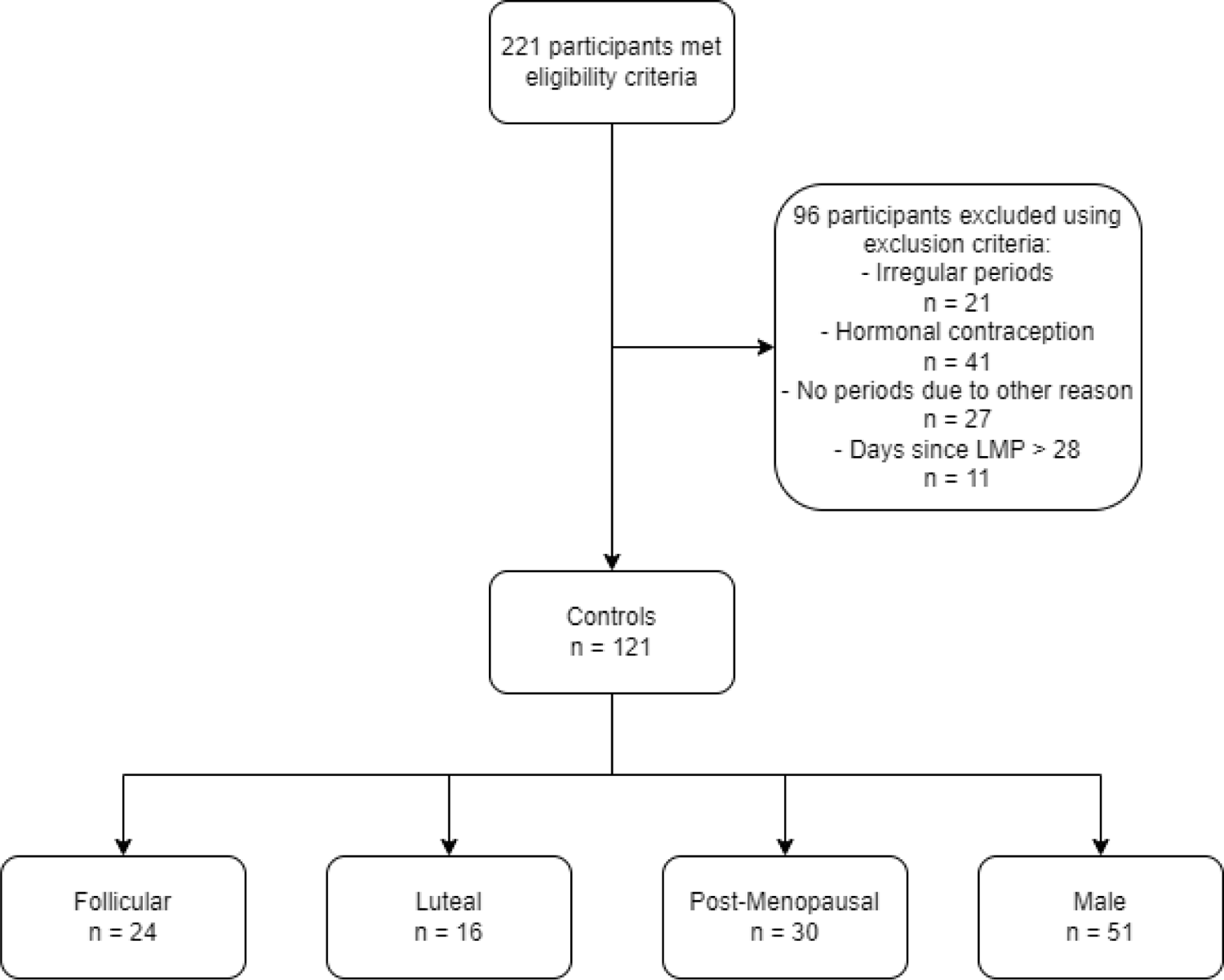
Participant eligibility and exclusion flowchart determining the number of participants in the final study population and menstrual cycle phase groups.

**Table 1:**
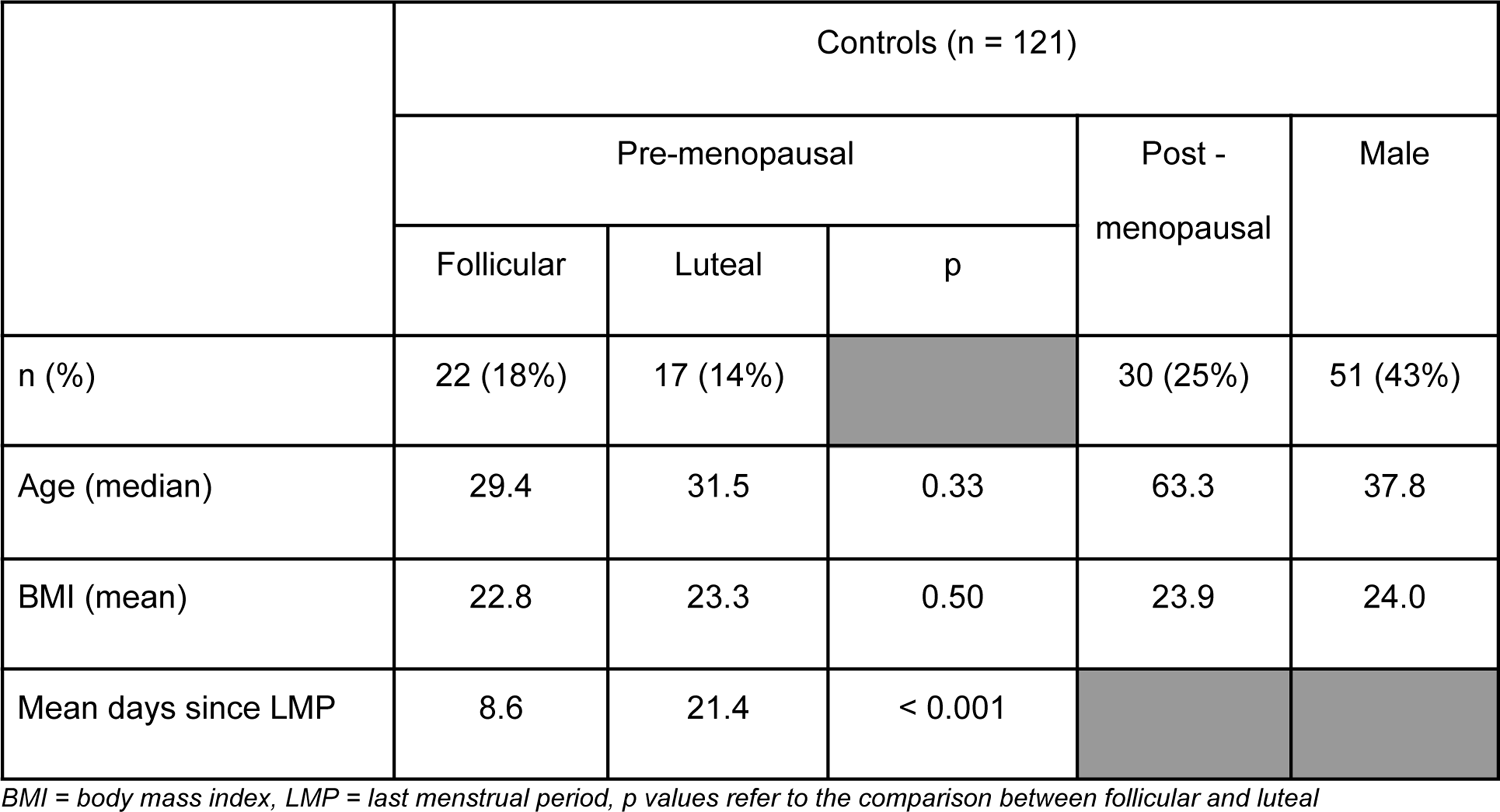
Overall Participant Demographics and Days Since Last Menstrual Period for Pre-menopausal Females.

|  | Controls (n = 121) |  |  |  |  |
| --- | --- | --- | --- | --- | --- |
|  | Pre-menopausal |  |  | Post -<br>menopausal | Male |
|  | Follicular | Luteal | p |  |  |
| n (%) | 22 (18%) | 17 (14%) |  | 30 (25%) | 51 (43%) |
| Age (median) | 29.4 | 31.5 | 0.33 | 63.3 | 37.8 |
| BMI (mean) | 22.8 | 23.3 | 0.50 | 23.9 | 24.0 |
| Mean days since LMP | 8.6 | 21.4 | < 0.001 |  |  |

BMI = body mass index, LMP = last menstrual period, p values refer to the comparison between follicular and luteal

### Gastric Electrophysiology Correlations

All averaged BSGM metrics were within the previously defined normal reference ranges for healthy control subjects across pre-menopausal females in both luteal and follicular phases, post-menopausal females and males (21). However, menstrual cycle phase and menopausal status were associated with significant electrophysiology variances within these reference ranges. As shown in **Figure 2A**, pre-menopausal females in the luteal phase had a significantly higher PGF than females in the follicular phase (mean 3.23 cpm, SD (0.15) vs. 2.92 cpm, SD (0.17), p < 0.001). Luteal phase gastric frequencies were also significantly higher than in males (mean 3.23 cpm, SD (0.15) vs 3.01 cpm, SD (0.2), p < 0.001).

**Figure 2:**
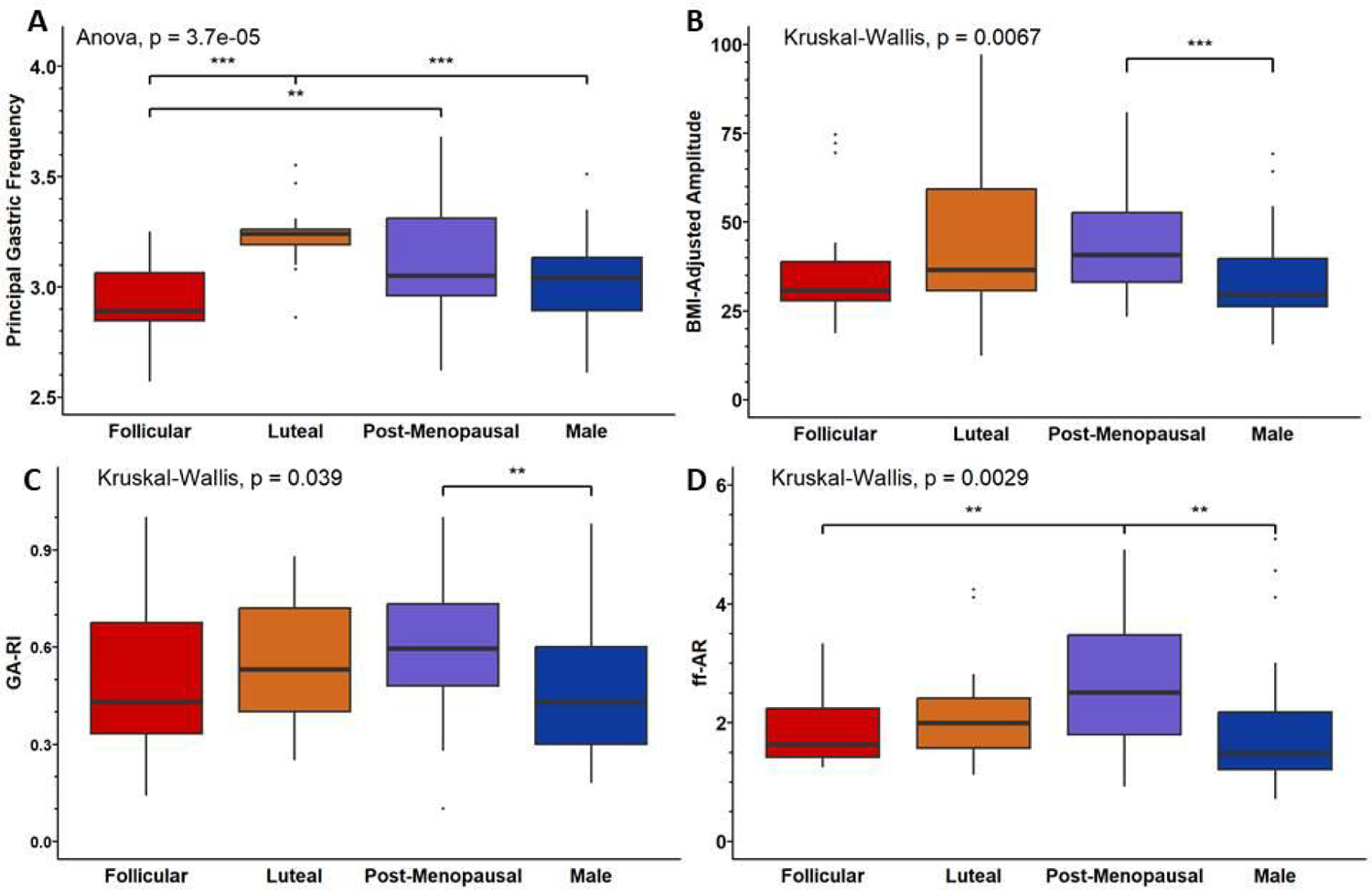
Box plot comparisons of overall BSGM metrics between pre-menopausal females in the follicular phase, pre-menopausal females in the luteal phase, post-menopausal females and males. A = Principal gastric frequency, B = BMI-Adjusted amplitude, C = Gastric Alimetry Rhythm Index (GA-RI), D = Fed:Fasted Amplitude Ratio

There were no differences in other metrics for pre-menopausal females in the luteal phase versus follicular phase, including BMI-adjusted amplitude (median 36.5 µV, (range 30.60 - 59.20) vs. 31.1 µV, (27.93 - 421.80), p = 0.39), GA-RI (median 0.53, (range 0.40 - 0.72) vs. 0.43, (0.33 - 0.68), p = 0.55) and ff-AR (median 1.99, (range 1.57 - 2.41) vs. 1.63, (1.42 - 2.24), p = 0.29). However, post-menopausal females showed a higher PGF than pre-menopausal females in the follicular phase (PGF mean 3.11 cpm, SD (0.24) vs. 2.92 cpm, SD (0.17), p = 0.003), as well as a higher ff-AR (median 2.51 (IQR 1.79 - 3.47) vs. 1.63 (1.42 - 2.24), p = 0.009).

Post-menopausal females additionally showed significant variations compared to men. As reported in **Figure 2B**, BMI-adjusted amplitude was higher in post-menopausal females compared to males (40.7 µV (33.02 - 52.58) vs. 29.6 µV (26.15 - 39.65), p < 0.001). Similarly, GA-RI was higher in post-menopausal females than males (0.60 (0.48 - 0.73) vs. 0.43 (0.30 - 0.60), p = 0.005) (**Figure 2C**), as was the ff-AR (2.51 (1.79 - 3.47) vs. 1.48 (1.21 - 2.17), p = 0.001) (**Figure 2D**).

### Meal Response Curves and Hourly Metrics

Meal response variations over the 4-hour post-prandial recording period were evaluated by generating summary plots combining all subjects’ data into a single spectral plot per subgroup (**Figure 3**) (13).

**Figure 3:**
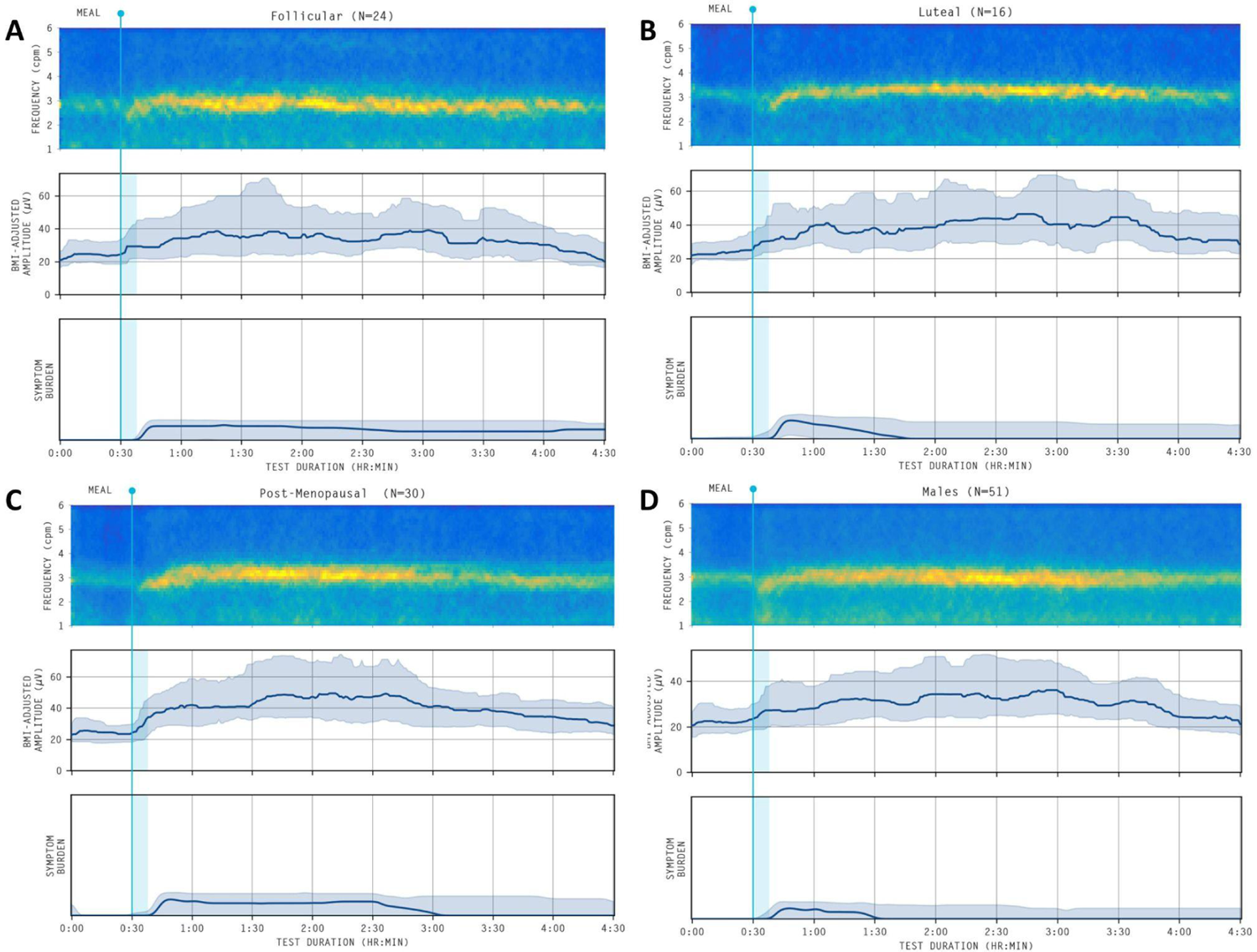
Average spectrograms summating Principal Gastric Frequency (top) and BMI-amplitude adjusted amplitude curves with shaded interquartile range (middle) and symptom burden curve across the duration of the Gastric Alimetry test. Meal initiation is indicated by a light blue vertical line at 30 minutes with a 10-minute consideration for consumption time (shaded). A = pre-menopausal females in the follicular phase, B = pre-menopausal females in the luteal phase, C = post-menopausal females, D = males.

Gastric meal response physiology was then systematically evaluated by comparing the pre-prandial half-hour and post-prandial hourly measurements for each metric (**Figure 4**). **Figure 4A** shows that differences in PGF between pre-menopausal females in the luteal and follicular phases were consistent throughout the entire meal response period, being elevated pre-prandially and at all post-prandial hours (all p < 0.01). **Figures 4B** and **4C** show significant differences between pre-menopausal females in the luteal phase and males 4 hours post-meal for BMI-adjusted amplitude (p = 0.03) and GA-RI (p = 0.04). BMI-adjusted amplitude was different between post-menopausal females and males at one-hour post-meal (p < 0.001), two hours post-meal (p = 0.002) and four hours post-meal (p = 0.003). **Figure 4C** also shows post-menopausal females consistently had a higher GA-RI, particularly in comparison to males, reaching statistical significance across the entire postprandial period (all p < 0.02).

**Figure 4:**
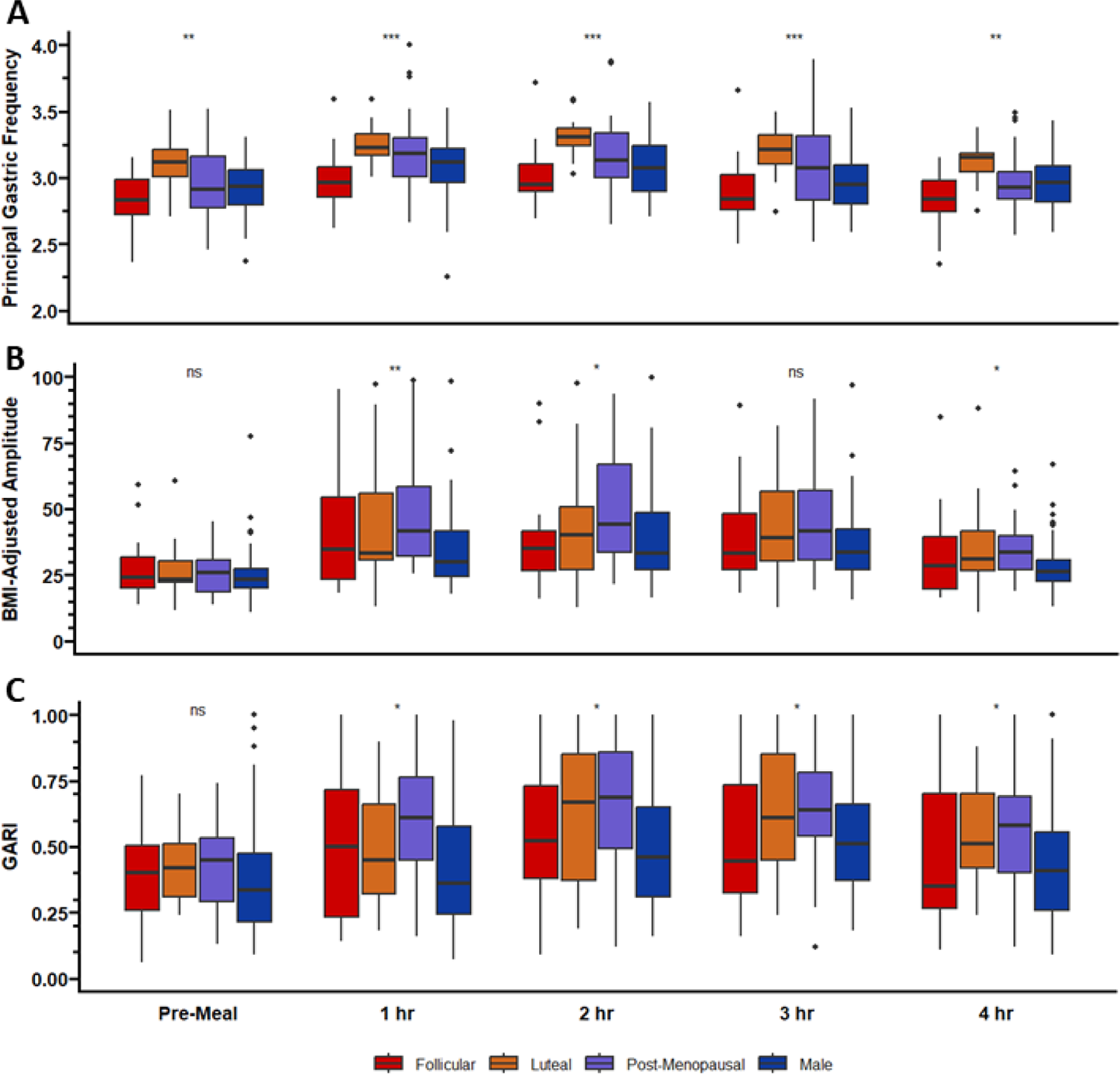
Box plot comparison of BSGM Metrics between menstrual cycle phase post-meal at one-hour intervals. A = Principal Gastric Frequency, B = BMI-Adjusted Amplitude, C = Gastric Alimetry Rhythm Index. 30 participants were not included in the BMI-adjusted amplitude analysis due to missing data.

### Symptom Burden

Participants experienced minimal symptoms regardless of menstrual phase or subgroup status, with no differences in total symptom burden measured continuously over the course of the Gastric Alimetry test (p = 0.52) (see symptom burden in **Figure 3** and **Figure 5A**). The predominant symptom was excessive fullness which was consistent across all groups (p = 0.35) (**Figure 5B**). The number of vomiting, belching and reflux events were insignificant between menstrual cycle phases and subgroup status.

**Figure 5:**
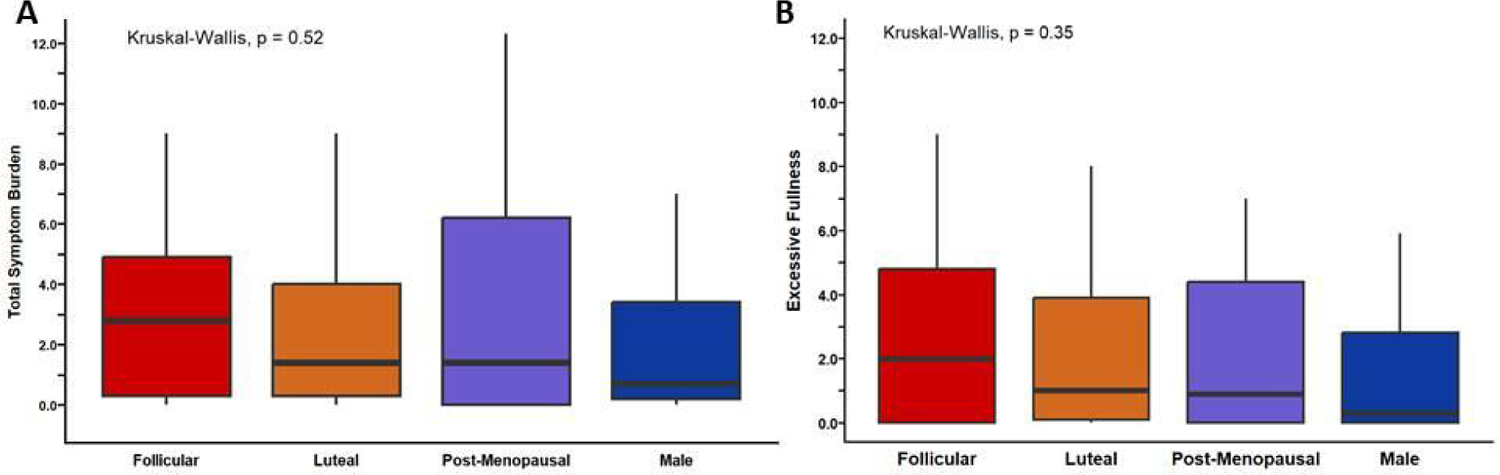
Box plot comparison of symptom burdens between menstrual cycle phases. A = Total Symptom Burden, B = Excessive Fullness

### Computational Model

In view of the significant frequency differences between the follicular and luteal menstrual phases, the gastric computational model was applied to estimate the resultant physiological consequences of this variability on gastric mixing and emptying. An illustrative frame from the computational model is shown in **Figure 6A**, with the full animations of both follicular (modelled at 2.94 cpm) and luteal phase (3.21 cpm) models available in the Supplementary Material. Overall, the models estimated that gastric mixing would increase approximately 8.1% (**Figure 6B**) and gastric emptying 5.3% (**Figure 6C**) in the luteal phase due to the frequency difference.

**Figure 6:**
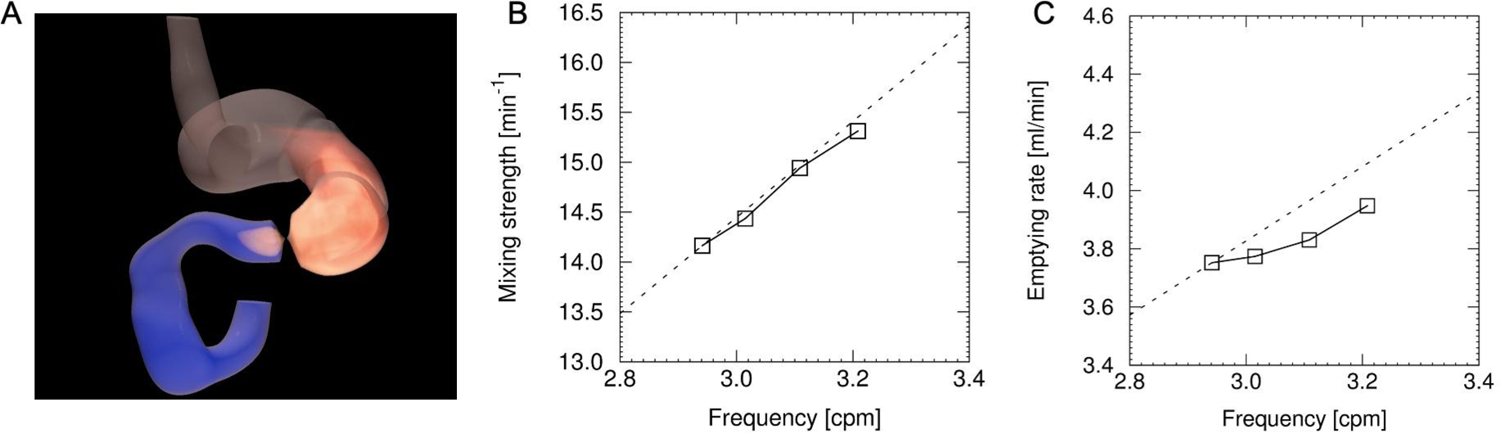
A = Illustrative frame of the gastric computational model (refer to Supplementary Material for animations). B, C = Gastric mixing strength and gastric emptying outcomes modelled across the experimentally-derived menstrual cycle frequency range. The linear dashed line represents the changes that would occur due to frequency changes alone, however, a restitution relationship exists such that gastric slow wave velocity falls as frequency increases (refer to (30)), giving rise to the result represented by the solid line.

## Discussion

This study has assessed the influence of the menstrual cycle and menopause on gastric electrophysiology. Significant novel variations in gastric function were observed in relation to menstrual cycling, with the luteal phase (when progesterone circulates systemically) characterised by a sustained increased Principal Gastric Frequency (PGF). Post-menopausal females also exhibited higher frequencies than females in the follicular phase and higher amplitudes and fed-fasted amplitude ratios (ff-AR) than men. These changes in gastric electrophysiology were not associated with symptomatic variation in this population of healthy subjects.

Changes to normal gastric function during the menstrual cycle have previously been documented, albeit using legacy electrogastrography (EGG) techniques, which have reduced resolution and lower reliability than modern high-resolution approaches (31). Most significantly, the administration of large doses of exogenous progesterone has been shown to provoke increased gastric dysrhythmia accompanied by increased frequency on EGG in healthy pre-menopausal controls (9). This frequency rise is consistent with the demonstrated luteal phase variability, although activity was exclusively of normal rhythm in this physiological study, as indicated by the GA-RI metric remaining within normative intervals (22). Two small, older studies using legacy EGG directly investigated the effect of the menstrual cycle, but these have shown no difference or subtle post-prandial frequency changes during the follicular phase (32,33). Notably, these previous studies only measured participants up to 20 days after the onset of menstruation, and periods of peak progesterone were amalgamated with the follicular phase or excluded altogether (32,33). Our study, using modern high-resolution recording techniques, overcomes the technological limitations of these previous studies by establishing that the luteal phase has a higher preprandial baseline frequency than the follicular phase (32). This finding also explains why female subjects in general show a higher average gastric slow wave frequency than males (22,32).

The changes observed in the luteal phases of this study are likely to be mediated by progesterone (9). After ovulation, the corpus luteum is formed, which releases progesterone to prepare the body for potential pregnancy during the luteal phase (34). Many human and animal model studies have evaluated the effects of progesterone on gastrointestinal transit and function, as recently reviewed by Coquoz et al (37). Results have been variable and sometimes conflicting, with some studies finding slower gastric emptying during the follicular phase (11,38). In this study, we extended our electrophysiological findings to predict the effects on gastric function using an established computational model of gastric mixing and emptying (26–28), finding that higher frequencies should result in increased gastric emptying in the luteal phase, even when the effects of velocity restitution are considered (30). These data are therefore consistent with the experimental study by Brennan et al who likewise found faster gastric emptying during the luteal phase in healthy subjects, in conjunction with increased appetite and energy intake during this phase (8). Progesterone elevation in the luteal phase increases basal core body temperature by 0.3 to 0.7 °C and is also associated with an increase in average heart rate by 3-5% compared to the follicular phase (35). The effects on heart rate were associated with a decrease in vagal activity and increased sympathetic dominance in the mid-luteal phase based on heart rate variability testing across the menstrual cycle (36). A convergence of physiological approaches therefore indicates that gastric function is accelerated in the luteal phase of the menstrual cycle, possibly through autonomic nervous system effects of progesterone, and that this may lead to increased gastric emptying.

A surprising finding in this study was the distinction in gastric electrophysiology between post-menopausal females and males, including higher amplitudes, more stable gastric rhythms (GA-RI), and higher fed:fasted amplitude ratios. Post-menopausal females and males have similar baseline progesterone and oestrogen levels (39,40), and menopausal hormone therapy (MHT) was not used amongst the post-menopausal group. The effect of MHT on gastric function is still unclear (32,41,42), while in animal models, testosterone has not been shown to have any effect on gastric motility (43). Comparative results between post-menopausal females and males should be interpreted with respect to the wider physiological context. Age may account for variations in the generation and transmission of gastric myoelectrical signals as interstitial cells of Cajal (ICC; “pacemaker” cells of the stomach) undergo age-related decline (44), although no significant age-specific effects were recently found on body surface recordings in a recent large normal population study (22). However, post-menopausal females are likely to have lower abdominal muscle:fat ratio, which could justify the higher amplitude and rhythm stability despite BMI adjustment of both metrics (21), although not explaining the higher ff-AR. It would be of interest to assess in future dedicated studies the responsible factors for these menopausal variations.

Inter-individual variability complicates menstrual cycle analysis. We recognise the limitations that arise when a 28-day menstrual cycle is assumed and menstrual cycle phase is allocated using a forward counting, day-based method, but this definition is widely used in the literature in the absence of sampling and hormonal quantification (45). The results produced have demonstrated robustness and high significance which is convincing for the influence of the menstrual cycle on the gastrointestinal system. The influence of female sex hormones on gastric electrophysiology can now therefore be assessed reliably and at scale in a non-invasive manner, which may be important in clinical trials where the menstrual cycle could otherwise be a confounding variable (8). We also acknowledge that many pre-menopausal females may have an alteration in their “baseline” or natural menstrual cycle pattern due to contraceptive use. Users of hormonal contraception were excluded from this study and remain an area for future exploration. Other pathologies, such as polycystic ovarian syndrome (PCOS), also alter sex steroids and may yield further insights (46).

In conclusion, this innovative study, using cutting-edge high-resolution gastric mapping and computational modelling techniques, found substantial variations in gastric activity with menstrual cycling and menopause. Most importantly, gastric slow wave frequency is significantly higher in the luteal versus follicular menstrual phase, which likely results in higher rates of gastric mixing and gastric emptying in the luteal phase. These findings indicate potential hormonal implications on gastric function that could influence symptoms in females with chronic gastroduodenal symptoms.

## Supporting information

Supplementary Animation 1A

Supplementary Animation 1B

## Acknowledgements

This research was conducted on behalf of the BSGM consortium. We thank India Wallace and Gen Johnson for their invaluable research assistance.

## Data Availability

Data used for analysis will be made available upon reasonable request, conditional on ethical approvals

## Supplementary Material

### Supplementary Animations

**Supplementary Figure 1 (at 2 x speed):**
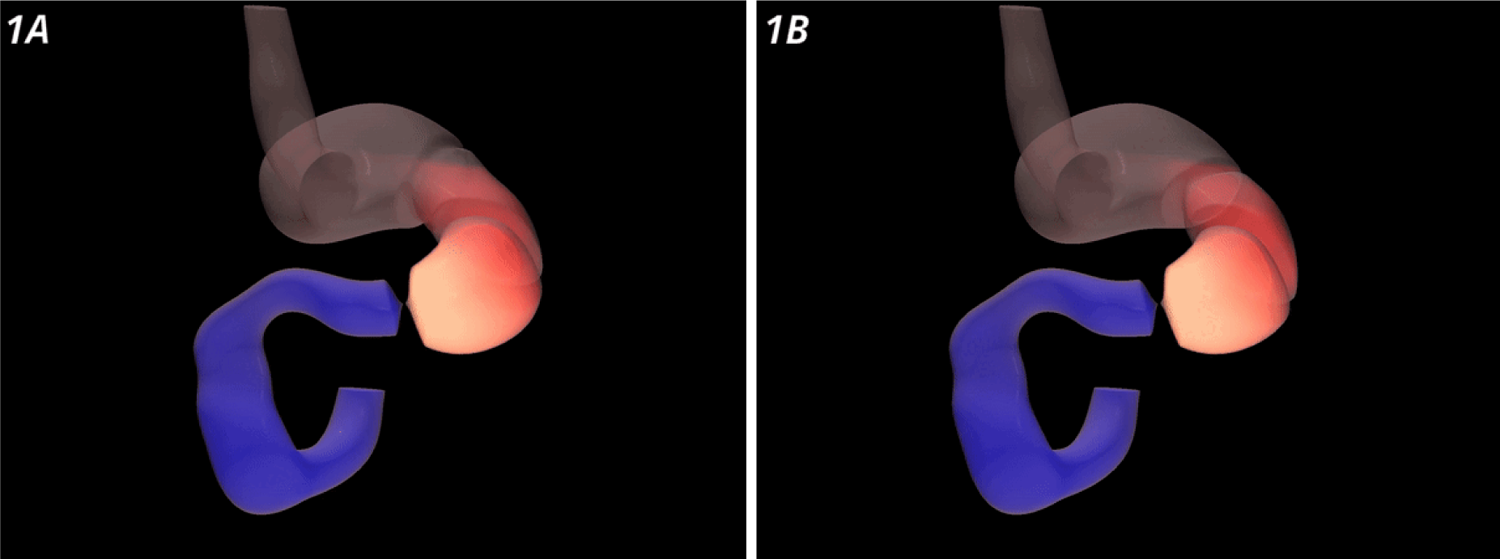
**1A** = Computational model of gastric fluid dynamics during the follicular phase of the menstrual cycle, programmed with frequency at 2.94 cpm. **1B** = computational model of gastric fluid dynamics during the luteal phase of the menstrual cycle, programmed with frequency at 3.21 cpm.

## References

1. Sperber AD, Bangdiwala SI, Drossman DA, Ghoshal UC, Simren M, Tack J, et al. Worldwide Prevalence and Burden of Functional Gastrointestinal Disorders, Results of Rome Foundation Global Study. Gastroenterology [Internet]. 2021 Jan;160(1):99–114.e3. Available from: 10.1053/j.gastro.2020.04.014

2. Black CJ, Drossman DA, Talley NJ, Ruddy J, Ford AC. Functional gastrointestinal disorders: advances in understanding and management. Lancet [Internet]. 2020 Nov 21;396(10263):1664–74. Available from: 10.1016/S0140-6736(20)32115-2

3. Jiang Y, Greenwood-Van Meerveld B, Johnson AC, Travagli RA. Role of estrogen and stress on the brain-gut axis. Am J Physiol Gastrointest Liver Physiol [Internet]. 2019 Aug 1;317(2):G203–9. Available from: 10.1152/ajpgi.00144.2019

4. Koloski NA, Talley NJ, Boyce PM. The impact of functional gastrointestinal disorders on quality of life. Am J Gastroenterol [Internet]. 2000 Jan 1;95(1):67–71. Available from: https://www.sciencedirect.com/science/article/pii/S0002927099007947

5. Talley NJ. Functional gastrointestinal disorders as a public health problem. Neurogastroenterol Motil [Internet]. 2008 May;20 Suppl 1:121–9. Available from: 10.1111/j.1365-2982.2008.01097.x

6. Zia JK, Heitkemper MM. Upper Gastrointestinal Tract Motility Disorders in Women, Gastroparesis, and Gastroesophageal Reflux Disease. Gastroenterol Clin North Am [Internet]. 2016 Jun;45(2):239–51. Available from: 10.1016/j.gtc.2016.02.003

7. Verrengia M, Sachdeva P, Gaughan J, Fisher RS, Parkman HP. Variation of symptoms during the menstrual cycle in female patients with gastroparesis. Neurogastroenterol Motil [Internet]. 2011 Jul;23(7):625–e254. Available from: 10.1111/j.1365-2982.2011.01681.x

8. Brennan IM, Feltrin KL, Nair NS, Hausken T, Little TJ, Gentilcore D, et al. Effects of the phases of the menstrual cycle on gastric emptying, glycemia, plasma GLP-1 and insulin, and energy intake in healthy lean women. Am J Physiol Gastrointest Liver Physiol [Internet]. 2009 Sep;297(3):G602–10. Available from: 10.1152/ajpgi.00051.2009

9. Walsh JW, Hasler WL, Nugent CE, Owyang C. Progesterone and estrogen are potential mediators of gastric slow-wave dysrhythmias in nausea of pregnancy. Am J Physiol [Internet]. 1996 Mar;270(3 Pt 1):G506–14. Available from: 10.1152/ajpgi.1996.270.3.G506

10. Wang F, Zheng TZ, Li W, Qu SY, He DY. Action of progesterone on contractile activity of isolated gastric strips in rats. World J Gastroenterol [Internet]. 2003 Apr;9(4):775–8. Available from: 10.3748/wjg.v9.i4.775

11. Gill RC, Murphy PD, Hooper HR, Bowes KL, Kingma YJ. Effect of the menstrual cycle on gastric emptying. Digestion [Internet]. 1987;36(3):168–74. Available from: 10.1159/000199414

12. Holtmann G. Functional Gastroduodenal Disorders. In: Practical Gastroenterology and Hepatology: Esophagus and Stomach [Internet]. Oxford, UK: Wiley-Blackwell; 2010. p. 434–41. Available from: https://onlinelibrary.wiley.com/doi/10.1002/9781444327311.ch55

13. Gharibans AA, Hayes TCL, Carson DA, Calder S, Varghese C, Du P, et al. A novel scalable electrode array and system for non-invasively assessing gastric function using flexible electronics. Neurogastroenterol Motil [Internet]. 2023 Feb;35(2):e14418. Available from: 10.1111/nmo.14418

14. Carson DA, O’Grady G, Du P, Gharibans AA, Andrews CN. Body surface mapping of the stomach: New directions for clinically evaluating gastric electrical activity. Neurogastroenterol Motil [Internet]. 2021 Mar;33(3):e14048. Available from: 10.1111/nmo.14048

15. O’Grady G, Varghese C, Schamberg G, Calder S, Du P, Xu W, et al. Principles and clinical methods of body surface gastric mapping: Technical review. Neurogastroenterol Motil [Internet]. 2023 Mar 29;e14556. Available from: 10.1111/nmo.14556

16. O’Grady G, Gharibans AA, Du P, Huizinga JD. The gastric conduction system in health and disease: a translational review. Am J Physiol Gastrointest Liver Physiol [Internet]. 2021 Nov 1;321(5):G527–42. Available from: 10.1152/ajpgi.00065.2021

17. Smout AJ, van der Schee EJ, Grashuis JL. What is measured in electrogastrography? Dig Dis Sci [Internet]. 1980 Mar;25(3):179–87. Available from: 10.1007/BF01308136

18. Stanghellini V, Chan FKL, Hasler WL, Malagelada JR, Suzuki H, Tack J, et al. Gastroduodenal Disorders. Gastroenterology [Internet]. 2016 May;150(6):1380–92. Available from: 10.1053/j.gastro.2016.02.011

19. Sebaratnam G, Karulkar N, Calder S, Woodhead JST, Keane C, Carson DA, et al. Standardized system and App for continuous patient symptom logging in gastroduodenal disorders: Design, implementation, and validation. Neurogastroenterol Motil [Internet]. 2022 Aug;34(8):e14331. Available from: 10.1111/nmo.14331

20. Gharibans AA, Calder S, Varghese C, Waite S, Schamberg G, Daker C, et al. Gastric dysfunction in patients with chronic nausea and vomiting syndromes defined by a novel non-invasive gastric mapping device [Internet]. Available from: 10.1101/2022.02.07.22270514

21. Schamberg G, Varghese C, Calder S, Waite S, Erickson J, O’Grady G, et al. Revised spectral metrics for body surface measurements of gastric electrophysiology. Neurogastroenterol Motil [Internet]. 2023 Mar;35(3):e14491. Available from: 10.1111/nmo.14491

22. Varghese C, Schamberg G, Calder S, Waite S, Carson D, Foong D, et al. Normative Values for Body Surface Gastric Mapping Evaluations of Gastric Motility Using Gastric Alimetry: Spectral Analysis. Am J Gastroenterol [Internet]. 2022 Dec 20; Available from: 10.14309/ajg.0000000000002077

23. Pitchers G & Elliot-Sale. Considerations-for-coaches-training-female-athletes. Professional Strength and Conditioning [Internet]. 2019; Available from: https://www.researchgate.net/publication/338126513

24. Waaseth M, Bakken K, Dumeaux V, Olsen KS, Rylander C, Figenschau Y, et al. Hormone replacement therapy use and plasma levels of sex hormones in the Norwegian Women and Cancer postgenome cohort - a cross-sectional analysis. BMC Womens Health [Internet]. 2008 Jan 14;8:1. Available from: 10.1186/1472-6874-8-1

25. Schmalenberger KM, Tauseef HA, Barone JC, Owens SA, Lieberman L, Jarczok MN, et al. How to study the menstrual cycle: Practical tools and recommendations. Psychoneuroendocrinology [Internet]. 2021 Jan;123:104895. Available from: 10.1016/j.psyneuen.2020.104895

26. Ishida S, Miyagawa T, O’Grady G, Cheng LK, Imai Y. Quantification of gastric emptying caused by impaired coordination of pyloric closure with antral contraction: a simulation study. J R Soc Interface [Internet]. 2019 Aug 30;16(157):20190266. Available from: 10.1098/rsif.2019.0266

27. Ebara R, Ishida S, Miyagawa T, Imai Y. Effects of peristaltic amplitude and frequency on gastric emptying and mixing: a simulation study. J R Soc Interface [Internet]. 2023 Jan;20(198):20220780. Available from: 10.1098/rsif.2022.0780

28. Berry R, Miyagawa T, Paskaranandavadivel N, Du P, Angeli TR, Trew ML, et al. Functional physiology of the human terminal antrum defined by high-resolution electrical mapping and computational modeling. Am J Physiol Gastrointest Liver Physiol [Internet]. 2016 Nov 1;311(5):G895–902. Available from: 10.1152/ajpgi.00255.2016

29. Pal A, Indireshkumar K, Schwizer W, Abrahamsson B, Fried M, Brasseur JG. Gastric flow and mixing studied using computer simulation. Proc Biol Sci [Internet]. 2004 Dec 22;271(1557):2587–94. Available from: 10.1098/rspb.2004.2886

30. Wang THH, Du P, Angeli TR, Paskaranandavadivel N, Erickson JC, Abell TL, et al. Relationships between gastric slow wave frequency, velocity, and extracellular amplitude studied by a joint experimental-theoretical approach. Neurogastroenterol Motil [Internet]. 2018 Jan;30(1). Available from: 10.1111/nmo.13152

31. Schamberg G, Calder S, Varghese C, Xu W, Wang WJ, Ho V, et al. Comparison of Gastric Alimetry^®^body surface gastric mapping versus electrogastrography spectral analysis [Internet]. bioRxiv. 2023. Available from: 10.1101/2023.06.05.23290993

32. Parkman HP, Harris AD, Miller MA, Fisher RS. Influence of age, gender, and menstrual cycle on the normal electrogastrogram. Am J Gastroenterol [Internet]. 1996 Jan;91(1):127–33. Available from: https://www.ncbi.nlm.nih.gov/pubmed/8561112

33. Tolj N, Luetić K, Schwarz D, Bilić A, Jurcić D, Gabrić M. The impact of age, sex, body mass index and menstrual cycle phase on gastric myoelectrical activity characteristics in a healthy Croatian population. Coll Antropol [Internet]. 2007 Dec;31(4):955–62. Available from: https://www.ncbi.nlm.nih.gov/pubmed/18217441

34. Oliver R, Pillarisetty LS. Anatomy, Abdomen and Pelvis, Ovary Corpus Luteum [Internet]. StatPearls Publishing; 2023 [cited 2023 Sep 11]. Available from: https://www.ncbi.nlm.nih.gov/books/NBK539704/

35. Baker FC, Siboza F, Fuller A. Temperature regulation in women: Effects of the menstrual cycle. Temperature (Austin) [Internet]. 2020 Mar 22;7(3):226–62. Available from: 10.1080/23328940.2020.1735927

36. de Zambotti M, Nicholas CL, Colrain IM, Trinder JA, Baker FC. Autonomic regulation across phases of the menstrual cycle and sleep stages in women with premenstrual syndrome and healthy controls. Psychoneuroendocrinology [Internet]. 2013 Nov;38(11):2618–27. Available from: 10.1016/j.psyneuen.2013.06.005

37. Coquoz A, Regli D, Stute P. Impact of progesterone on the gastrointestinal tract: a comprehensive literature review. Climacteric [Internet]. 2022 Aug;25(4):337–61. Available from: 10.1080/13697137.2022.2033203

38. Wald A, Van Thiel DH, Hoechstetter L, Gavaler JS, Egler KM, Verm R, et al. Gastrointestinal transit: the effect of the menstrual cycle. Gastroenterology [Internet]. 1981 Jun;80(6):1497–500. Available from: https://www.ncbi.nlm.nih.gov/pubmed/7227774

39. Sheffield JS, Wendel GD Jr, McIntire DD, Norgard MV. The effect of progesterone levels and pregnancy on HIV-1 coreceptor expression. Reprod Sci [Internet]. 2009 Jan;16(1):20–31. Available from: 10.1177/1933719108325510

40. Abraham GE, Swerdloff R, Tulchinsky D, Odell WD. Radioimmunoassay of plasma progesterone. J Clin Endocrinol Metab [Internet]. 1971 May;32(5):619–24. Available from: 10.1210/jcem-32-5-619

41. Hutson WR, Roehrkasse RL, Wald A. Influence of gender and menopause on gastric emptying and motility. Gastroenterology [Internet]. 1989 Jan;96(1):11–7. Available from: 10.1016/0016-5085(89)90758-0

42. Gonenne J, Esfandyari T, Camilleri M, Burton DD, Stephens DA, Baxter KL, et al. Effect of female sex hormone supplementation and withdrawal on gastrointestinal and colonic transit in postmenopausal women. Neurogastroenterol Motil [Internet]. 2006 Oct;18(10):911–8. Available from: 10.1111/j.1365-2982.2006.00808.x

43. Chen TS, Doong ML, Chang FY, Lee SD, Wang PS. Effects of sex steroid hormones on gastric emptying and gastrointestinal transit in rats. Am J Physiol [Internet]. 1995 Jan;268(1 Pt 1):G171–6. Available from: 10.1152/ajpgi.1995.268.1.G171

44. Gomez-Pinilla PJ, Gibbons SJ, Sarr MG, Kendrick ML, Shen KR, Cima RR, et al. Changes in interstitial cells of cajal with age in the human stomach and colon. Neurogastroenterol Motil [Internet]. 2011 Jan;23(1):36–44. Available from: 10.1111/j.1365-2982.2010.01590.x

45. Schmalenberger KM, Eisenlohr-Moul TA, Würth L, Schneider E, Thayer JF, Ditzen B, et al. A Systematic Review and Meta-Analysis of Within-Person Changes in Cardiac Vagal Activity across the Menstrual Cycle: Implications for Female Health and Future Studies. J Clin Med Res [Internet]. 2019 Nov 12;8(11). Available from: 10.3390/jcm8111946

46. Elliott-Sale KJ, Minahan CL, de Jonge XAKJ, Ackerman KE, Sipilä S, Constantini NW, et al. Methodological Considerations for Studies in Sport and Exercise Science with Women as Participants: A Working Guide for Standards of Practice for Research on Women. Sports Med [Internet]. 2021 May;51(5):843–61. Available from: 10.1007/s40279-021-01435-8

